# Improvised Water Procurement in Wilderness Medicine: A Comparative Review of Yield, Energy Cost, and Field Suitability

**DOI:** 10.1101/2025.07.10.25331307

**Authors:** Inam S. Ahmed

## Abstract

**Introduction:** Access to potable water remains a fundamental need in wilderness and disaster medicine. Improvised hydration techniques—such as solar stills, transpiration bags, and rain catchment—are widely taught but poorly compared. This review evaluates six low-resource water procurement methods for survival relevance.

**Methods:** A structured narrative review was conducted across PubMed, Google Scholar, and grey-literature sources from January 1990 to March 2025. Included studies reported water yield, labor time, disinfection potential, or energy cost. A modified Newcastle–Ottawa Scale assessed study quality (max = 8). Comparative energy-return-on-hydration (EROH) was calculated using MET estimates.

**Results:** Twenty-seven studies (17 field, 10 lab) from 10 countries were included (NOS median 5, IQR 4–6). Rain catchment yielded 5–10 L/event with minimal labor and no energy cost. Direct filtration produced >10 L/day when surface water was available. Boiling achieved complete pathogen removal but required fuel and time. Solar stills produced <0.6 L/day and demanded 375–500 kcal per build. Dew and transpiration methods yielded <1 L/day and required favorable humidity or sunlight. Solar stills were the least energy-efficient method. Table 1 summarizes comparative metrics; Figure 2 presents a decision-support algorithm.

**Conclusions:** Solar stills, though historically emphasized in survival curricula, offer poor yield and an unfavorable energy profile. Modern tools like filters, catchment tarps, and chemical disinfection should be prioritized in both practice and training. Wilderness educators and responders should re-align survival instruction with evidence-based hydration strategies.

## INTRODUCTION

Access to potable water is a universal necessity, but in wilderness, disaster, and conflict zones, obtaining clean drinking water becomes a major operational and survival challenge. Dehydration can degrade cognitive function, reduce physical endurance, and increase the risk of hypothermia, heat illness, and renal injury—conditions frequently encountered in austere settings.^1,2,3^ Under stressful conditions like survival scenarios, even mild dehydration has been shown to impair thermoregulation and decision-making within hours.^4,5,6^ For these reasons, reliable access to safe water is prioritized in wilderness medicine protocols and disaster response operations globally.

Improvised water procurement techniques—those that do not require pre-manufactured devices —are commonly taught in survival training programs. These include solar stills, transpiration bags, rain catchment systems, dew cloth collection, boiling, and makeshift filters. Many of these techniques are prominently featured in military field manuals (e.g., FM 21-76), civilian survival handbooks, and instructional media.^7,8^ However, these techniques vary widely in their effectiveness, complexity, and regional suitability.

Historically, the solar still achieved near-iconic status in survival manuals and military curricula, often portrayed as a universal fallback method. Promoted as a passive, gear-free way to distill water from soil or vegetation, it has long been cited in military, scout, and survivalist literature. ^7,8,9^ Yet recent field experiments,^10^ metabolic energy analyses,^11^ and critical reviews by the Wilderness Medical Society^12^ have raised serious doubts about its practicality, highlighting its low yield, intensive labor demands, and environmental limitations. In contrast, modern survival kits now often include compact filtration systems, water-treatment tablets, and collapsible catchment tarps that are far more effective.

This review examines six low-resource water procurement methods using standardized criteria: water yield, labor and energy cost, disinfection potential, and environmental suitability. The goal is to inform evidence-based protocols for wilderness providers, rescue teams, and field educators.

## METHODS

Search Strategy: A structured narrative review was conducted across PubMed, Google Scholar, and grey-literature sources including government and NGO repositories (e.g., USDA, CDC, EPA, WHO, FEMA). Searches were performed from 1 January 1990 to 31 March 2025 using Boolean-modified keywords and phrases: “ solar still,” “ rain catchment,” “ wilderness water,” “ transpiration bag,” “ dew collection,” “ emergency hydration,” “ portable filter,” and “ survival water yield.” Secondary snowball citation tracking was used to identify additional relevant studies.

Inclusion: We included field-based and laboratory-controlled studies that measured the water yield (in liters per day) or pathogen-removal capacity of improvised water procurement methods. Studies had to report at least one quantitative datapoint related to water output, microbial reduction, or filtration efficiency. Acceptable interventions included solar stills, transpiration bags, rain catchment systems, dew cloths, boiling, and portable filtration devices.

Exclusion: We excluded studies that focused solely on desalination technologies designed for boats or industrial use, models without real-world validation, or conceptual frameworks lacking empirical data. Our aim was to retain studies that could directly inform low-resource, field-relevant practices.

Data Extraction & Quality: One reviewer independently extracted key metrics from each study, including daily water yield (liters per 24 h), labor requirements (in minutes), and disinfection potential. To ensure consistency, data were cross-checked against reported values in the original publications and re-tabulated into a standardized comparison matrix. Study quality was assessed using a modified Newcastle–Ottawa Scale (max score = 8), which considered selection methods, real-world relevance, outcome measurement, and potential bias. Although no second reviewer was available, an internal calibration process using three pilot studies ensured consistency of scoring criteria across all included works.

Energy-Cost Analysis: Labor kcal expenditure for solar-still digging was calculated using Ainsworth’s Compendium MET of 8.5 for “ shoveling sand.” ^13^

Figures: Figure 1 presents a PRISMA flow diagram summarizing the study identification and screening process. Figure 2 provides a decision-support algorithm to help field clinicians and survival educators choose the most suitable water procurement technique based on environment, equipment availability, and contamination risk.

## RESULTS

Study Characteristics: Twenty-seven studies (17 field-based, 10 laboratory-based) conducted in 10 countries met inclusion criteria (Figure 1). Study quality was moderate overall, with a median Newcastle–Ottawa Scale (NOS) score of 5 (IQR 4–6). Most field studies simulated real-world survival scenarios, while lab-based studies focused on controlled comparisons of device efficacy. Common outcome measures included water yield (liters/day), labor time (minutes), microbial log reduction, and environmental constraints.

Yield and Labor Burden: Water yield and labor effort varied significantly across techniques (Table 1). Rain catchment systems consistently delivered the highest volume (median: 5–10 L/ event) with minimal labor (<15 min). Direct filtration methods yielded >10 L/day when surface water was available and required <5 minutes per use. Boiling was effective but labor- and fuel-intensive, requiring 15 minutes per liter and fuel access. Solar stills produced only 0.3–0.6 L/day and demanded 45–60 minutes of construction effort, while transpiration bags yielded 0.3–1 L/ day. Dew cloths yielded the least (0.1–0.3 L/day), though they required minimal physical effort.

**TABLE 1.** Comparative Performance of Low-Resource Water-Procurement Methods.

| Method | Equipment Needed | Median Yield (L day<sup>-1</sup>) | Setup/Labor (min) | Pathogen Removal | Best Environment |
| --- | --- | --- | --- | --- | --- |
| Rain catchment | Tarp/poncho + container | 5–10 † per rainfall <sup>11,12</sup> | 15 | None | Humid, rainy |
| Direct filtration | Hollow-fiber filter | >10 ‡ (source-limited) | <5 | High (bacteria/protozoa) | Any surface water |
| Boiling | Pot + heat source | Unlimited (fuel-limited) | 15 per L | Complete <sup>5</sup> | Fuel-rich basecamps |
| Solar still | Plastic sheet + cup | 0.3–0.6 | 90 | Complete | Sunny, arid, moist |
| Transpiration bag | Clear plastic bag | 0.3–1 | 5 | None | Vegetated sunlight |
| Dew cloth | Cloth/mesh | 0.1–0.3 | 10 | None | Clear, humid nights |
| †Volume per event;<br>‡Flow-rate dependent. |  |  |  |  |  |

Energy Efficiency: Using a MET value of 8.5 for “ shoveling sand,” solar still construction required ∼500 kcal, making it the least energy-efficient method. In contrast, rain catchment provided >5 L of water for <40 kcal investment. When assessed as energy return on hydration (EROH), solar stills scored lowest.

Pathogen Mitigation: Boiling reliably achieves >4-log *E. coli* and >3-log *Giardia* reductions, as supported by WHO water treatment benchmarks.^15^ Solar stills may reduce pathogen load through phase separation, though direct log-reduction data remain limited; however, solar disinfection using transparent-bottle models (SODIS) has demonstrated >4-log *E. coli* reductions under high UV conditions.^16^

Environmental Suitability: Technique effectiveness was context-dependent. Rain catchment was optimal in tropical, coastal, or temperate zones. Direct filtration was viable anywhere surface water existed. Boiling was most effective at basecamps with fuel. Transpiration bags and dew cloths worked in sunny or humid climates respectively. Solar stills were marginally useful in hot, arid regions when plastic sheeting was available and no other water sources were present.

## DISCUSSION

This review affirms the wide performance disparity across improvised water procurement methods and supports reevaluating the longstanding instructional emphasis on solar stills. While their appeal as gear-free survival tools is well-established in manuals and handbooks, real-world field data reveal serious limitations in their utility. Their water yield is minimal (0.3–0.6 L/day), the construction effort is calorically expensive (∼500 kcal per setup), and their dependency on specific environmental conditions—sunlight, moist soil, and clear plastic—renders them unreliable for most scenarios.

In contrast, rain catchment systems provided the most favorable yield-to-effort ratio, requiring minimal labor (<15 min setup) and yielding >5 L per rainfall event. These systems benefit from scalability, broad environmental applicability, and zero ongoing energy cost. Direct filtration methods, particularly those meeting NSF P231 certification, offered the greatest versatility and ease of use, particularly in zones with available surface water. Boiling remains the most effective technique for pathogen inactivation, but its limitations include fuel dependency, time, and potential safety concerns in unstable field environments.

Notably, this review emphasizes the energetic inefficiency of solar stills through energy-return-on-hydration (EROH) analysis. In calorically constrained survival conditions, techniques that require high energy expenditure for low yield are not just impractical—they may be detrimental to survival outcomes.

The pathogen mitigation profile of methods was also highly variable. While boiling and NSF-validated filtration were robust, solar stills and improvised methods like transpiration and dew collection offered little inherent disinfection unless paired with post-treatment steps. This finding reinforces the need for multi-step or hybrid approaches in water procurement strategies where safety is critical.

Environmental suitability further stratified the utility of each technique. Rain catchment and filtration performed best in humid, temperate, or coastal zones. Boiling was favorable at basecamps with reliable fuel sources. Transpiration bags and dew cloths were situationally effective in sunny vegetative zones or humid nights, respectively. Solar stills, by contrast, remained viable only in hot, arid environments where no surface water exists and plastic sheeting is on hand—a rare convergence in survival scenarios.

Altogether, these findings challenge outdated doctrines in survival education and highlight the need for curriculum reform. Modern field kits that include compact filters, water tablets, or even collapsible catch systems offer dramatically better returns in both yield and safety. Wilderness medicine instructors, disaster responders, and outdoor educators should be encouraged to align their recommendations with this evolving evidence base.

Limitations: Heterogeneity in field methodology, limited long-term data, and variability in terrain and weather reduce the comparability of studies. Most yield measurements derive from short-term or single-event trials, limiting inference on sustainability, reliability across climates, and user fatigue over multi-day survival scenarios. In addition, many studies were conducted under near-optimal laboratory or fair-weather field conditions, which may not reflect real-world adversity such as overcast skies, fuel scarcity, altitude, or cold exposure. Few studies systematically assessed technique performance under operational fatigue or logistical constraint. Furthermore, because this review was conducted by a single reviewer, despite internal calibration steps, the possibility of data extraction or scoring bias cannot be fully excluded.

## CONCLUSIONS

Evidence supports de-prioritizing solar stills in modern wilderness curricula. Instructors should emphasize more effective techniques such as rapid rainwater harvesting, surface-water filtration, and fuel-efficient boiling. Although solar stills were once widely promoted—particularly in Cold War-era military manuals, Boy Scouts handbooks, and FEMA survival training—they are no longer the preferred method due to their inefficiency, environmental dependence, and energy cost. Their persistence in outdated training models reflects legacy thinking more than functional utility.

With the availability of compact water filters, chemical disinfectants, ultralight stoves, and collapsible catchment systems, modern survival kits now offer reliable, multi-use solutions that far surpass solar stills in both practicality and safety. Rain catchment systems require minimal labor and offer high yield, especially in temperate or tropical environments. Direct filtration systems perform well in nearly any terrain where surface water exists and now routinely meet NSF P231 certification standards. Boiling, while fuel-dependent, remains the gold standard for pathogen elimination when conditions permit.

Solar stills, by contrast, provide less than 0.6 liters per day under optimal conditions and demand considerable labor—approximately 375–500 kcal per construction. When energy return on hydration (EROH) is considered, solar stills often produce a net deficit in calorically limited survival settings. Additionally, the availability of the required materials—clear plastic sheeting, a collection vessel, and suitable soil—is not guaranteed in most realistic wilderness scenarios.

Instructional emphasis should shift toward tools and techniques with proven reliability across environments and minimal dependency on environmental variables. Field educators and clinical wilderness instructors must revisit the hierarchy of water procurement techniques they teach. Just as outdated practices like tourniquet avoidance have been corrected in trauma care, so too should ineffective water strategies be phased out of field medicine curricula.

Going forward, wilderness medicine training programs should revise their core modules to reflect this evidence-based prioritization. Practical training should include not only filtration and catchment setup, but also methods for field-testing water safety, improvising disinfection under limited resources, and recognizing signs of dehydration and hyponatremia. Further studies should evaluate combined or hybrid approaches, such as boiling filtered water or pairing catchment with SODIS treatment, to optimize safety and redundancy.

While the solar still remains an interesting demonstration of distillation principles and could serve as a fallback in desert survival scenarios where no alternative exists, its role should be strictly confined to academic discussion and historical review—not frontline teaching. Survival doctrine must evolve with data. This review provides a framework to modernize hydration education and better prepare field personnel, disaster responders, and backcountry clinicians for the realities of water procurement in austere environments.

## Data Availability

All data produced in the present study are available upon reasonable request to the authors

## ACKNOWLEDGMENTS

The author thanks Wilderness Medical Society peers for critical feedback.

## FINANCIAL/MATERIAL SUPPORT

None.

## FIGURE LEGENDS

Figure 1. PRISMA flow diagram: 218 records identified; 174 screened; 27 studies included.

Figure 2. Decision algorithm for selecting water-procurement technique based on environment, equipment, and contamination risk.

