## Supplementary figures and images for "Improvised Water Procurement in Wilderness Medicine: A Comparative Review of Yield, Energy Cost, and Field Suitability"

### Figure 1

Figure 2. Decision Algorithm for Water Procurement Method Selection

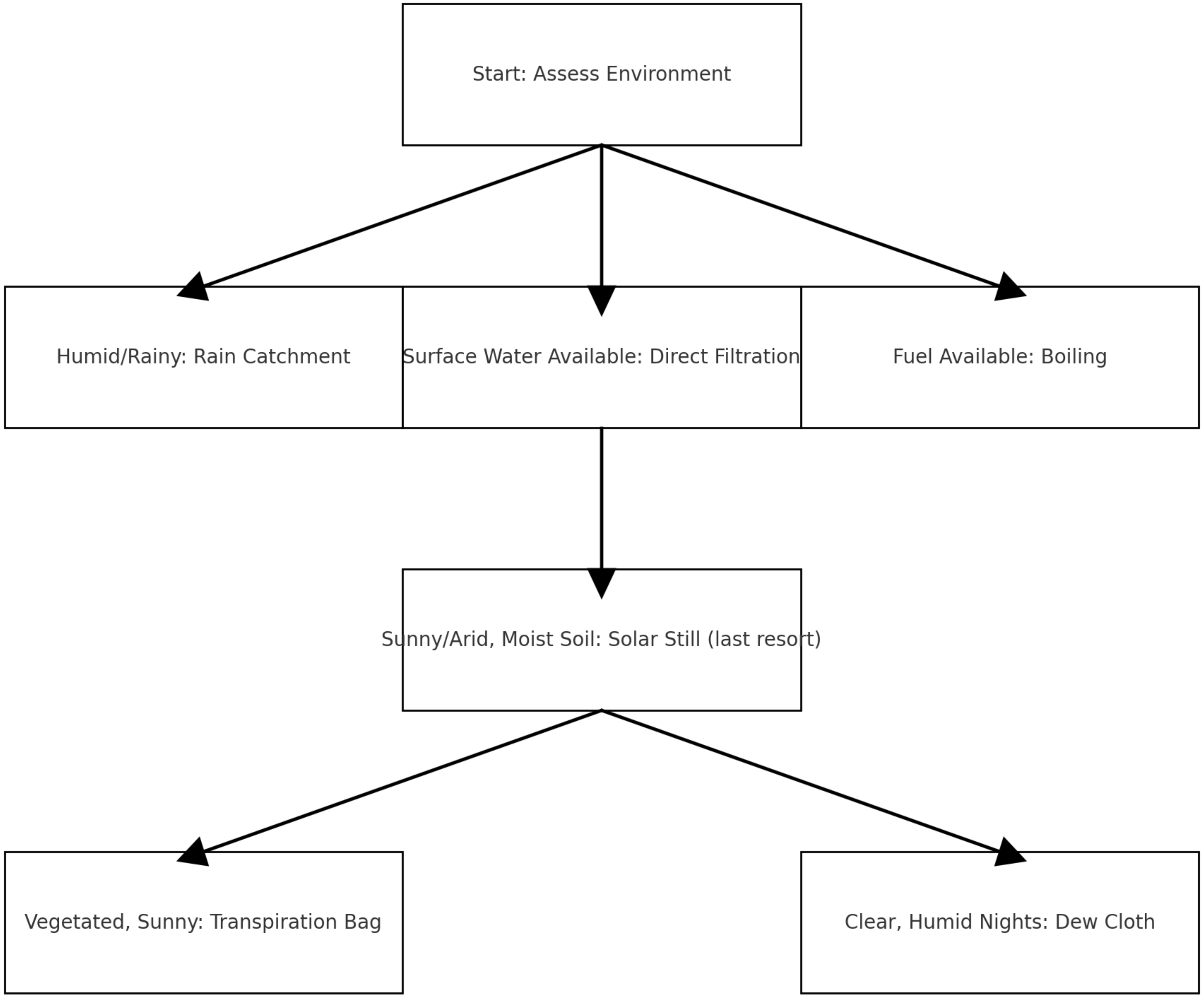

### Figure 2

Figure 1. PRISMA Flow Diagram

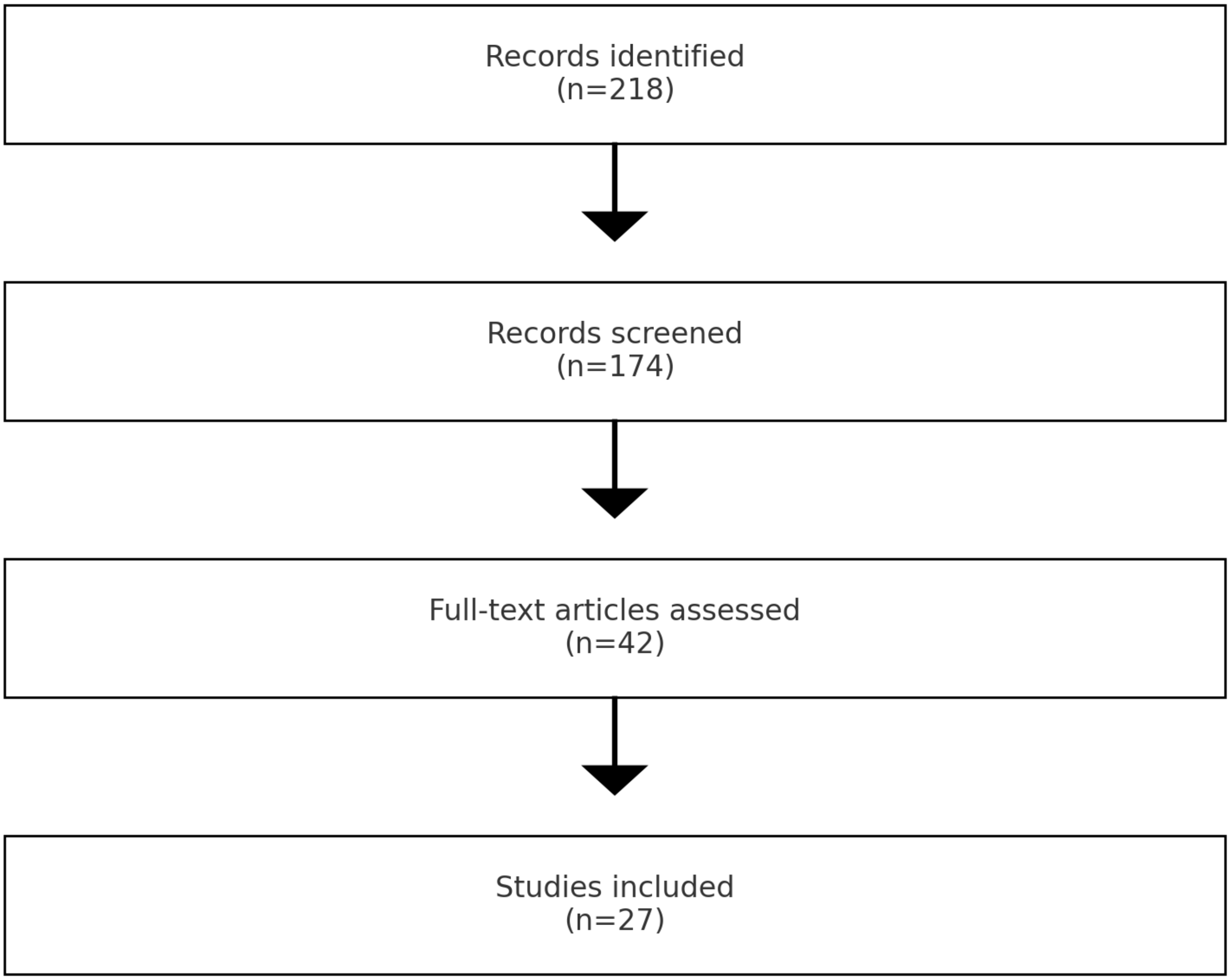
